# Patch nonwear accounts for more than half of apparent nonadherence in a digital medicine system

**DOI:** 10.64898/2026.09.21.26363541

**Authors:** Andrew Jin Soo Byun, Diana S. Chen, Haruka Notsu, Sunnie Li, Victoria Lisowski, Erlend Lane, Sean Ryan, John Torous

## Abstract

Digital-medicine systems can objectively confirm medication ingestion, but interpretation of a no-detection day depends on whether the wearable receiver was in contact with the body. We analyzed 997 person-days from 20 outpatients prescribed aripiprazole through Abilify MyCite by integrating ingestion records with patch-contact data; 17 participants also contributed smartphone sensing. Of 322 days without a detected ingestion, 53.7% had insufficient patch wear. Counting all no-detection days as nonadherent yielded an apparent nonadherence rate of 32.3%, whereas conditioning on adequate wear yielded 18.1%. Because adherence during patch-off periods is unobserved, the rate is bounded between 14.9% and 32.3%. These findings support a three-state outcome that distinguishes detected ingestion, no ingestion despite adequate wear, and patch-off or indeterminate days. Among 744 patch-worn days, detected ingestion was more likely on days that more closely matched each participant’s usual activity pattern (odds ratio 1.52, 95% credible interval 1.14–2.03) and when participants spent more time at home around the habitual dose period (1.31, 1.04–1.64). These behavioral associations were exploratory and characterize the context of medication-taking rather than causal effects. Overall, device wear was integral to interpretation of the adherence outcome, and passive behavioral measures were most interpretable after observation status was established.

**AUTHOR SUMMARY:** Digital tools can help researchers measure whether medications are taken in everyday life, but these tools are only useful when we know whether they were actually able to observe the behavior of interest. We studied 20 adults using a digital aripiprazole system in which an ingestible sensor communicates with a wearable patch. We found that more than half of days with no detected ingestion occurred when the patch was not worn long enough for a dose to have been observed. As a result, simply treating every day without a detected signal as medication nonadherence substantially overestimated apparent nonadherence.

We therefore propose distinguishing three types of days: days when an ingestion is detected, days when no ingestion is detected despite adequate patch wear, and days when medication-taking cannot be determined because the patch was not adequately worn. We also explored whether smartphone-derived measures of daily behavior were related to medication-taking. More typical daily activity patterns and being at home around the usual dosing time were associated with detected ingestion, but these findings were exploratory and did not support reliable day-ahead prediction. Our findings highlight the importance of establishing whether digital health data are observable and interpretable before drawing clinical conclusions.

## INTRODUCTION

Medication nonadherence contributes to relapse, rehospitalization, and functional decline across psychiatric disorders [1–3]. Our sample includes outpatients with major depressive disorder prescribed adjunctive aripiprazole, but the measurement problem addressed here arises from the device and applies across diagnoses. In schizophrenia, nonadherence estimates approach 56%, and missed doses and symptom worsening may reinforce one another over time [4]. Hospitalization risk can rise within days after an interrupted prescription refill [5], motivating adherence measures with finer temporal resolution than monthly pharmacy records. Yet self-report, clinician judgment, pill counts, and pharmacy records tend to overestimate adherence, poorly resolve individual missed doses, and correlate only modestly with objective ingestion [6,7].

Digital-medicine systems provide a more direct, day-resolved measure. Abilify MyCite combines aripiprazole with an ingestible sensor that emits a signal on contact with gastric fluid. A wearable adhesive patch detects the signal, records ingestion time, and relays the event to a smartphone application; the system received US Food and Drug Administration approval in 2017 [8,9]. Independent evaluations report ingestion-detection rates of 94% to 98% [10–12]. The central measurement limitation is dependence on patch wear. When no ingestion is detected, the dose may have been missed or the patch may not have been in contact with the skin. Interpretation therefore requires distinguishing adequately observed no-ingestion from absence of observation.

After observability is established, smartphone digital phenotyping can characterize the daily context of medication-taking. Accelerometry, screen interaction, GPS, and EMA capture activity, location, and engagement in everyday life [13–17]. Medication-taking may be less likely when behavior departs from an individual’s usual routine or when the individual is away from the usual dosing location. Prior work has linked passive sensing to symptoms, relapse, and engagement [19–22], but adherence studies have generally relied on self-report, pill counts, or outcome labels that do not account for device wear. Misclassified outcomes can produce spurious associations or apparent predictive performance, which makes within-person analysis particularly important [18].

Our group previously showed that digital-medicine and smartphone-phenotyping data can be collected together in this population, but confirmed ingestion was available on a median of only about 15% of expected study days, limiting analysis [24]. The present study uses the same class of system with the mindLAMP platform and applies an analytic sequence in which observation status is defined before behavioral context is evaluated.

The primary analysis quantified the effect of patch-blind classification on day-level adherence and represented unobserved days using a three-state outcome. Secondary analyses evaluated whether, among adequately observed days, detected ingestion was associated with within-person variation in daily routine and location, and whether similar patterns were observed for patch-off status or EMA nonresponse. A prospective analysis assessed whether passive sensing predicted a subsequent wear-supported no-ingestion day. This analytic sequence separates adherence ascertainment from behavioral association and prediction.

## RESULTS

### Sample and objective adherence

Twenty participants contributed objective ingestion and patch-contact data across 997 person-days (median 36.5 per participant, range 16–106). Dosing was predominantly once daily, with a single detected ingestion on 98.7% of ingestion days. The 17-participant behavioral sample provided 744 patch-worn days, including 128 wear-supported no-ingestion days, for a wear-conditioned adherence rate of 82.8%; 15 participants contributed all four data sources. Adherence varied substantially across participants, with P(adherent | worn) ranging from 0.18 to 0.98 (median 0.77), and three participants contributed 41% of all wear-supported no-ingestion days. Accelerometer-derived features met the prespecified coverage threshold on 72% of patch-window days, GPS data were available on 71%, and a mindLAMP EMA response was available on 54.1%. Patch wear, sensor coverage, and EMA response declined over time, whereas adherence conditional on patch wear remained comparatively stable, with only a mild weekend decrease.

### Patch nonwear accounts for more than half of no-detection days

Across all 20 participants, the three-state classification identified 675 adherent days (67.7%), 149 wear-supported no-ingestion days (14.9%), and 173 patch-off or indeterminate days (17.4%). Of 322 person-days without a detected ingestion, 173 (53.7%) had insufficient rather than adequate wear. Counting every no-detection day as nonadherent yielded an apparent nonadherence rate of 32.3%, whereas conditioning on adequate wear yielded 18.1%. Neither directly estimates the true missed-dose rate. Because patch-off days are unobserved, the rate is bounded between 14.9% if all patch-off days were adherent and 32.3% if none were; 18.1% assumes that adherence on unobserved days resembles adherence on observed days. Two observations weakly support that assumption: adherence conditional on wear remained stable as wear declined (Fig 1E), and patch-off days did not differ behaviorally from adherent days. Patch contact was strongly bimodal, with most days showing either nearly continuous or little to no contact, and the correction was stable across wear thresholds of 1 to 12 hours. Fig 1 summarizes the study overview and wear-based correction.

**Fig 1.**
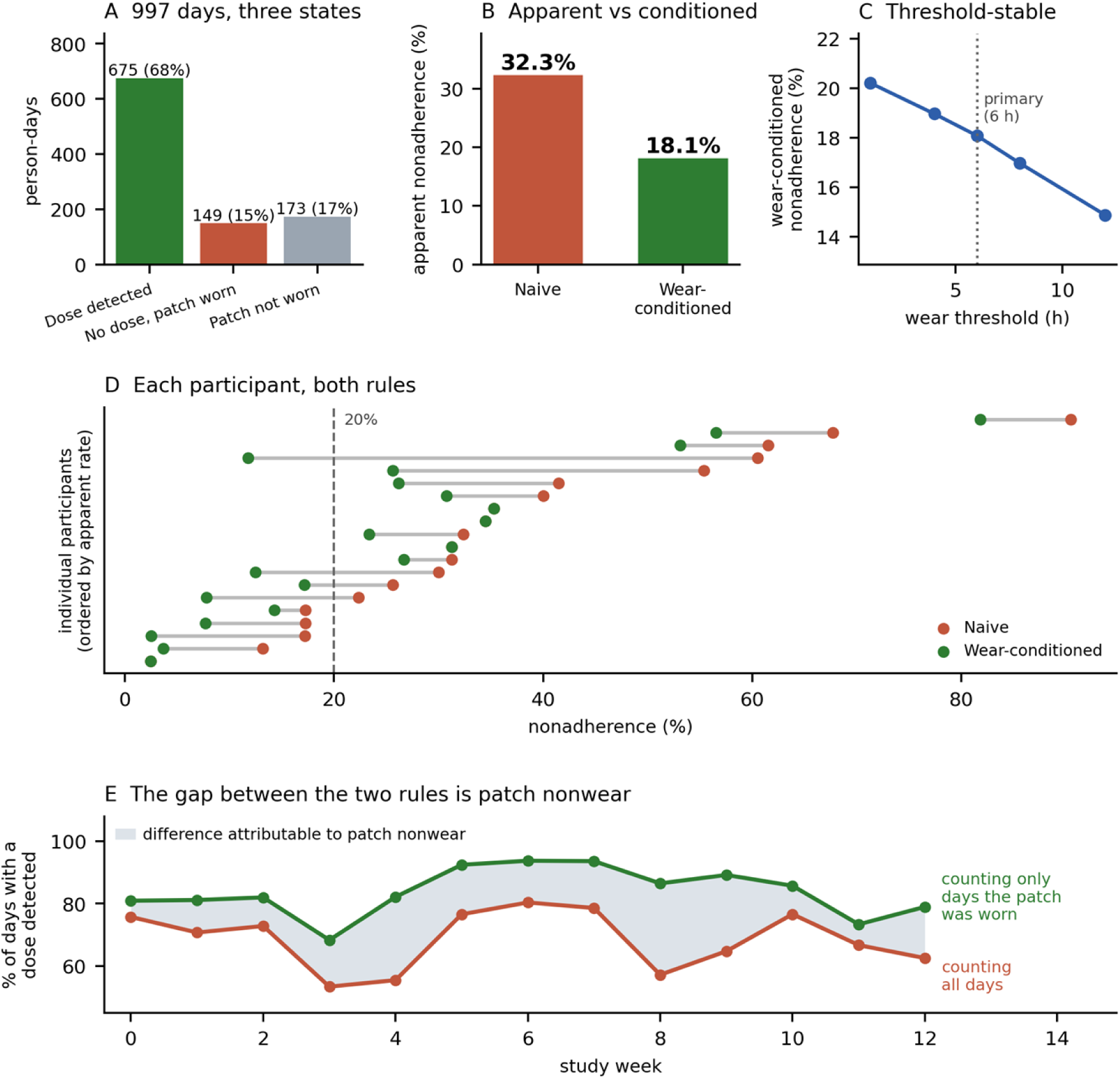
Patch wear and the day-level adherence outcome. (A) Each of the 997 person-days falls into one of three states: a dose was detected (675, 67.7%); no dose was detected although the patch was worn for at least 6 hours (149, 14.9%); or the patch was not worn long enough for a dose to have been observed (173, 17.4%). (B) Counting every day without a detected dose as a missed dose gives an apparent nonadherence rate of 32.3%; counting only days on which a dose could have been observed gives 18.1%. Neither is the true rate — because patch-off days are unobserved rather than known, the truth lies between 14.9% and 32.3%. (C) The same comparison repeated for wear thresholds from 1 to 12 hours, showing the correction does not depend on the 6-hour cut-off. (D) The two rules applied to each participant separately, ordered by apparent rate. Conditioning on wear moves four of 20 participants across a 20% threshold and moves one from 60.5% to 11.8%, so it changes which individuals look nonadherent, not only the group average. (E) The proportion of days with a detected dose in each study week, computed both ways. Counting all days (red) the proportion falls over the study; counting only days the patch was worn (green) it does not. The shaded gap between them is the share of days on which a dose could not have been observed, and it widens in the weeks when patch wear was lowest.

### Activity-rhythm regularity is associated with same-day adherence

Activity-rhythm regularity showed the clearest within-person association with adherence. A one-SD shift toward the participant’s usual activity pattern was associated with 52% higher odds of same-day adherence (posterior OR 1.52, 95% credible interval 1.14 to 2.03; posterior probability of a positive association, 99.8%). Given an adherence rate of 82.8% on patch-worn days, this effect corresponds to an increase in predicted probability from approximately 83% to 88%. The GEE sensitivity analysis was directionally consistent (OR 1.37, 95% CI 1.06 to 1.78; p=0.017), although the association did not survive correction across 12 features (q=0.12; Fig 2A) and is considered exploratory. Estimates were stable across sensitivity analyses (1.32–1.64), leave-one-participant-out analyses (1.39–1.63), and adjustment for patch-wear hours (OR 1.66, 1.24–2.23). No other primary feature had a credible interval excluding 1 (Fig 2).

**Fig 2.**
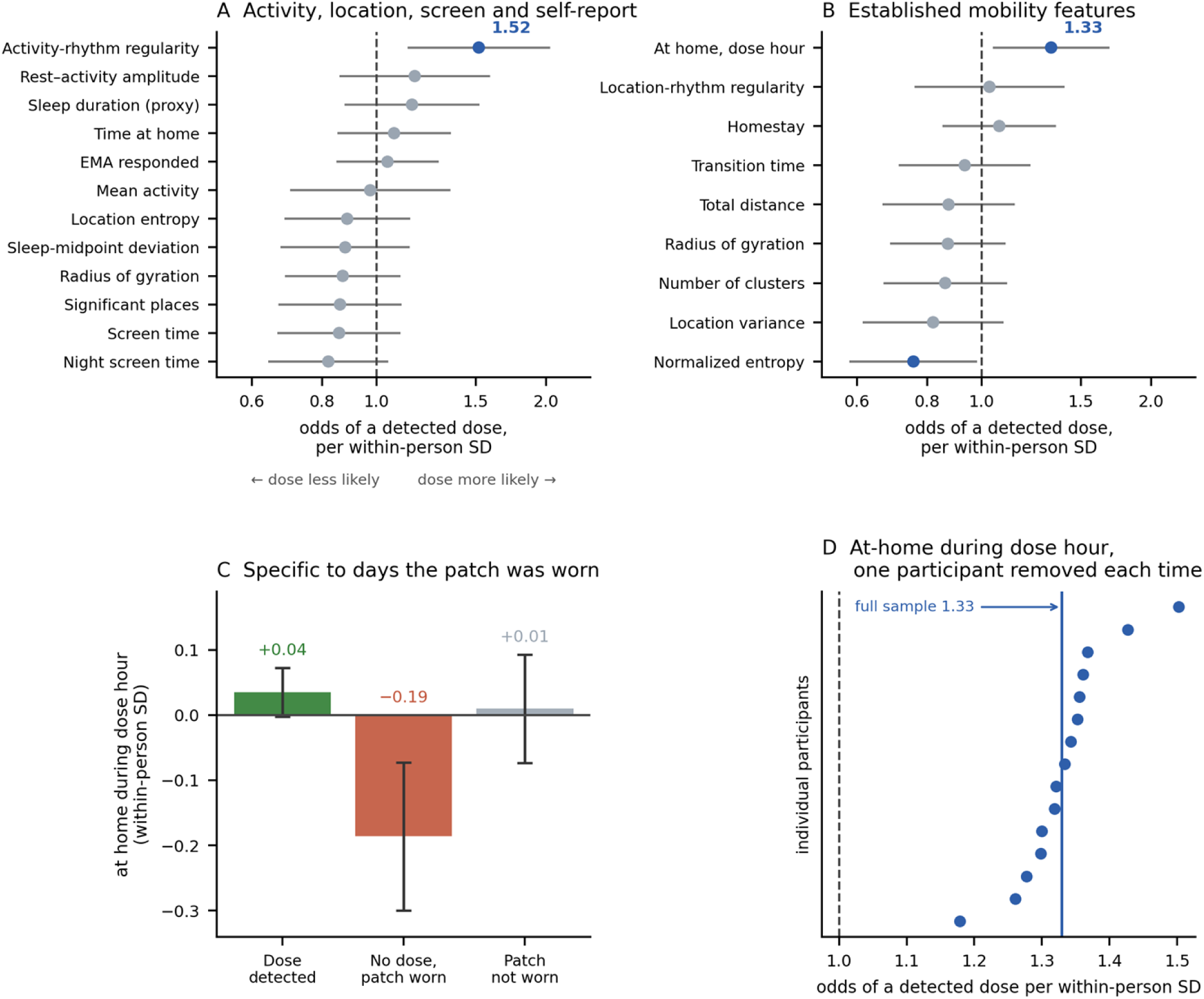
What daily behavior tells us about whether a dose was taken. Each feature is centered on that participant’s own average, so an odds ratio above 1 means a dose was more likely to be detected on days when the feature ran above that person’s usual level; the estimates describe day-to-day variation within people, not differences between them. Bayesian mixed-effects logistic regression on the 744 days the patch was worn, adjusted for weekend, study week, sensor coverage and survey response; blue marks intervals excluding 1. (A) The 12 prespecified features. Only activity-rhythm regularity — how closely a day resembled that person’s own typical activity pattern — excludes 1 (OR 1.52), and it does not survive correction for testing 12 features (q=0.12). (B) Daily mobility features as defined in prior digital-phenotyping work (Saeb et al. 2015); none survived correction (all q≥0.60). The two measures above the dotted line are not standard features but were built for this study: location-rhythm regularity repeats the activity-rhythm test using location instead of movement and is null (OR 1.03), so the routine association does not reappear in a second sensor; at-home during the habitual dose hour repeats the peri-dose test with a stricter, leave-one-day-out window and reproduces it (OR 1.33). Derivations for every feature are in Supplementary Table S1. (C) How much of the habitual dose hour was spent at home, by day state. The reduction appears only on days when the patch was worn and no dose was recorded, not on patch-off days. (D) The dose-hour estimate recomputed 15 times, each time leaving one participant out; all remain above 1, so no single participant drives it.

Time at home around the habitual dose time showed a second, weaker association (posterior OR 1.31, 95% credible interval 1.04 to 1.64; GEE p=0.087, q=0.27). A stricter measure of time at home during the single habitual dose hour, with that hour estimated leave-one-day-out, produced a similar estimate (OR 1.33, 1.05 to 1.69; Fig 2B) and retained its direction when any single participant was removed (Fig 2D). Movement and screen use in the same window, as well as all features from the preceding four hours, showed no association. The at-home association was confined to the dosing window and therefore characterizes the contemporaneous dosing context rather than a preceding signal. It may also be partly definitional because participants who dose at home are necessarily at home when ingestion is detected.

### Peri-dose at-home time is associated with adherence

The activity-rhythm association did not replicate across sensor modality. Among seven established daily mobility features (location variance, normalized entropy, homestay, transition time, total distance, number of clusters, and radius of gyration), none was associated with adherence after correction (all q≥0.60; Fig 2B). Normalized entropy had a credible interval excluding 1 (OR 0.76, 0.58 to 0.98) but did not survive correction. The location analogue of activity-rhythm regularity was null (OR 1.03, 95% credible interval 0.76 to 1.40), and its direction changed in leave-one-participant-out analyses. Time at home during the habitual dose hour was the only location measure that showed a consistent association with adherence, reproducing the peri-dose result under a stricter definition and retaining the same direction when any single participant was removed (OR range 1.18 to 1.50; Fig 2D). On patch-off days, at-home time remained near each participant’s average rather than being reduced (Fig 2C), so the reduction was confined to days with adequate patch wear and no detected ingestion.

### Patch-off days do not show the same descriptive activity-rhythm pattern

The same behavioral features were tested against patch-off status and EMA nonresponse to assess whether the routine-related findings reflected broader study disengagement. No feature remained associated with either outcome after correction for multiple comparisons (all q≥0.31). Descriptively, activity-rhythm regularity was near each participant’s personal norm when a dose was detected (within-person z=0.04) and when the patch was not worn (z=−0.02), whereas days with adequate wear but no detected ingestion showed greater deviation from the participant-specific routine (z=−0.16). Time at home during the habitual dose hour showed the same ordering (z=+0.04 adherent, −0.19 worn with no ingestion, +0.01 patch-off). This descriptive pattern was compatible with variation across ingestion-defined states rather than uniform study disengagement. However, the outcomes were not formally compared, and null findings for patch-off status and EMA nonresponse do not establish specificity.

### Passive sensing does not support within-person next-day prediction

Passive data through the end of day t−1 did not reliably predict nonadherence on day t. In pooled out-of-fold predictions, the multimodal model reached an AUROC of 0.686 (95% CI 0.557 to 0.777), compared with 0.642 for covariates alone and 0.65 for a model using only each participant’s first-week adherence rate (PR-AUC 0.34, 0.32, and 0.33; base rate 0.171; Fig 3). The pooled discrimination primarily reflected between-participant differences in nonadherence rates rather than within-participant day-level prediction: median within-participant AUROC was 0.49 (IQR 0.38 to 0.67), and a permutation test preserving each participant’s miss rate and temporal autocorrelation showed that the pooled result was compatible with the absence of day-level signal (p=0.126). Predicted probabilities were also overconfident (calibration slope 0.34, partly reflecting class-weighted training) and should not be interpreted as day-level risk estimates.

**Fig 3.**
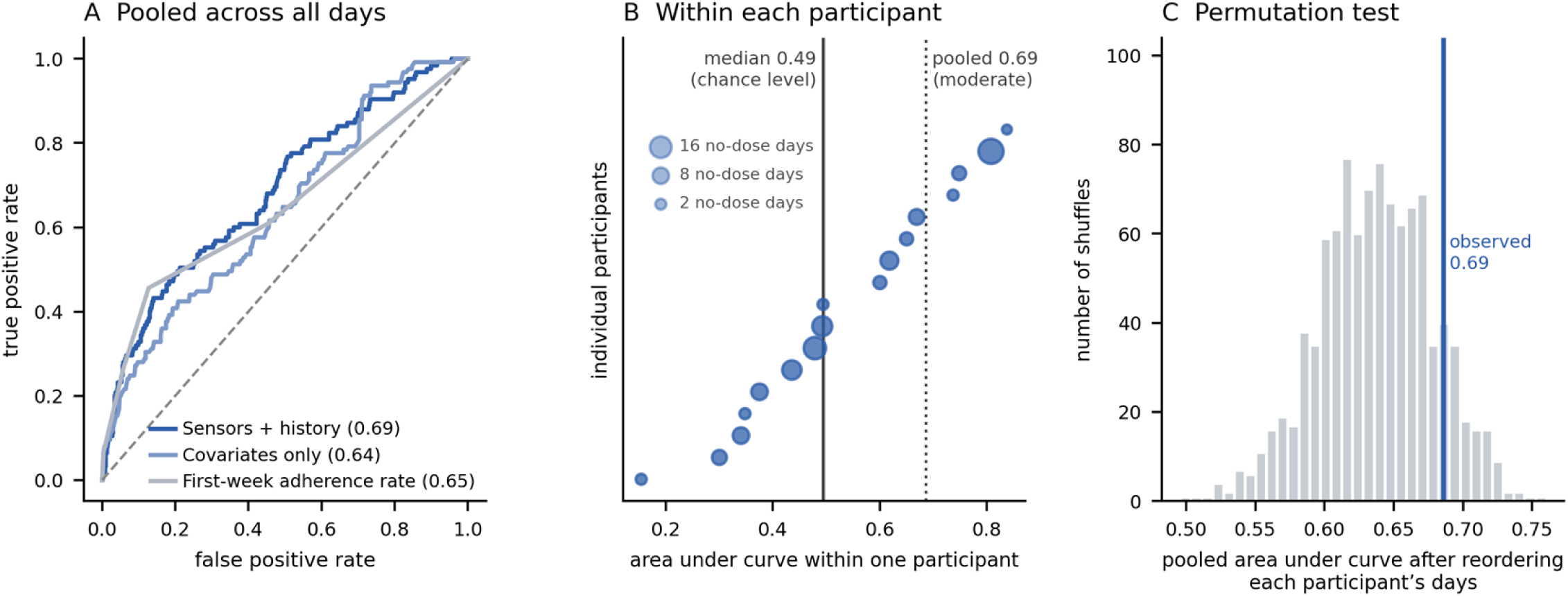
Passive sensing does not identify which days a dose will be missed. Models were fitted on all participants but one and evaluated on the held-out participant, so all values are out-of-sample. (A) Discrimination pooled across every participant-day for three models: sensor features plus adherence history, participant covariates only, and a model using nothing but each participant’s adherence rate during their first week. Areas under the curve of 0.69, 0.64 and 0.65 indicate that sensor data added little beyond knowing how often a participant had already missed doses. (B) The same sensor model evaluated within each participant separately. Circle size is proportional to the number of no-dose days that participant contributed. The median across participants was 0.49, which is chance performance — within a given participant the model could not distinguish days on which a dose was missed from days on which it was taken. The pooled value of 0.69, conventionally described as moderate discrimination, therefore reflects stable differences between participants in how often they miss doses rather than any day-level ability. (C) The pooled analysis repeated 1,000 times, each time reordering one participant’s days among themselves. This removes any relationship between a day’s behavior and that day’s outcome while leaving each participant’s overall rate of missed doses unchanged. Thirteen per cent of these reorderings reached the observed pooled value, so the pooled result falls within the range expected when no day-level signal is present.

### Longer-than-usual nighttime device inactivity is associated with same-day ingestion

Nighttime rest periods estimated from smartphone accelerometry measure sustained device inactivity rather than validated sleep, and 11% of well-observed nights exceeded 12 hours—durations implausible for sleep but not for a stationary phone. Longer-than-usual rest-period time was associated with higher odds of a same-day detected dose (posterior OR per within-person SD 1.42, 95% credible interval 1.05–1.90; GEE OR 1.23, within-family q=0.003). The association was graded across the full range rather than confined to extreme nights: adherence rose monotonically across quintiles of within-person rest deviation; a quadratic term was not supported (p=0.81); adding a >12-hour indicator left the linear trend intact (+0.052 per hour, p=0.016) while the indicator itself was not supported (OR 1.39, p=0.29); and the trend remained present when nights longer than 12 hours were excluded (+0.050 per hour, p=0.034). The long nights were among the best-observed measurements in the dataset (n=88; 95% with complete 24-hour coverage; mean efficiency 0.875) and occurred in 16 of 17 participants, supporting their interpretation as valid periods of prolonged device inactivity rather than recording failures. The signal was not explained by time at home, weekend, overall movement, screen time, or patch-wear hours, but it overlapped with activity-rhythm regularity: in mutual adjustment, rest-period time was attenuated to OR 1.16 (p=0.22), whereas activity-rhythm regularity remained at 1.36 (p=0.06). We therefore interpret longer-than-usual device rest as a further expression of a consolidated, routine day rather than an independent effect, and not as evidence about sleep physiology in either direction.

### EMA completion reflects between-participant rather than within-participant differences

Adherence was higher on days with a mindLAMP EMA response than on days without one (88% versus 76%), but this reflected stable between-participant differences (OR 1.70, 95% credible interval 1.38–2.10). Within participants, responding to the survey on a given day was not associated with same-day ingestion (OR 1.05, 0.85–1.29). No individual EMA symptom measure remained associated with adherence after correction for multiple comparisons. The strongest was self-reported anxiety, with higher anxiety associated with lower odds of same-day adherence (OR 0.76, 95% credible interval 0.58 to 0.99; q=0.12), a preliminary result similar in evidential strength to the activity-rhythm finding.

## DISCUSSION

The principal finding is that digital-medicine adherence estimates depend on observation status. More than half of no-detection days lacked adequate patch wear, and classifying all no-detection days as nonadherent increased apparent nonadherence from 18.1% among adequately worn days to 32.3%. A three-state outcome therefore distinguishes detected ingestion, adequately observed no-ingestion, and periods in which ingestion status cannot be determined. Within adequately observed days, detected ingestion also showed exploratory associations with similarity to each participant’s usual activity pattern and with time spent at home around the habitual dose period. These analyses place behavioral context downstream of adherence ascertainment and reduce the risk that missing observation is interpreted as medication nonadherence.

The patch-wear finding has implications beyond this dataset. In adherence technologies that depend on a worn receiver, absence of a recorded event conflates medication-taking with the ability to observe medication-taking. Treating device-off days as missed doses can distort group adherence rates and participant-level classification, as illustrated in Fig 1, and may also affect intervention estimates or model evaluation. The wear-conditioned rate does not represent a definitive missed-dose estimate because it assumes that adherence on unobserved days resembles adherence on observed days. A transparent reporting framework should therefore include the three-state outcome, the wear-conditioned rate, bounds implied by patch-off days, the proportion of no-detection days attributable to nonwear, and the adjudication rules. Contemporaneous self-report, unavailable here, could further help adjudicate no-detection days. This framework addresses the data-completeness limitation in our earlier digital-pill pilot, where confirmed ingestion was available on a median of about 15% of expected study days [24], and the broader concern that validation of an ingestion sensor does not resolve the classification of ambiguous no-detection days [23].

The behavioral analyses supported a narrower interpretation than a general routine-disruption effect. Detected ingestion was more likely on days that resembled each participant’s usual activity profile, whereas an analogous location-rhythm measure did not show the same association. Time at home was associated with ingestion specifically around the habitual dose period, not during the preceding hours. These findings are consistent with two distinct contextual correlates of medication-taking: consistency of the overall daily activity pattern and the immediate dosing environment. The at-home association could reflect environmental cues or access to medication, but it is also compatible with reverse description or confounding by symptoms and daily obligations. The results are consistent with prior work linking sleep and circadian disruption to adherence in schizophrenia [27,28] and with digital-phenotyping studies in which within-person variation is more informative than between-person differences [18].

Prospective analyses further distinguished contemporaneous association from prediction. The pooled multimodal model showed moderate discrimination, but within-participant AUROC was at chance, indicating that much of the pooled signal reflected stable between-participant differences in adherence rather than day-specific variation within individuals. Thus, the behavioral associations observed here should be interpreted as descriptive correlates of medication-taking context rather than as day-ahead risk markers. Larger prospective studies are needed to determine whether stable daily events or location-specific cues can support adherence interventions.

The rest-period analysis illustrates a construct-validity requirement rather than a data artifact. Smartphone accelerometry measures sustained device inactivity, and nights exceeding conventional sleep durations were among the best-observed measurements of that construct rather than implausible estimates; a plausibility filter defined by sleep physiology is not an appropriate validity check for a phone-derived measure, and restricting the exposure to its middle range attenuates a graded association mechanically. Because the association with ingestion was graded across the full range and largely shared with activity-rhythm regularity, we interpret it within the routine-consistency pattern and caution against relabeling device inactivity as sleep. Measurement error nonetheless affected both sides of the analysis: adherence status was ambiguous when the patch was not worn, and a phone-derived predictor was initially misread as a sleep measure. Both argue for explicit evaluation of observation status and construct validity before interpreting digital-phenotyping associations.

Several limitations constrain interpretation. The primary behavioral sample included only 17 participants, and three accounted for 41% of the wear-supported no-ingestion days. The data support day-level within-person analyses of moderate effects but not stable between-person comparisons, subgroup analyses, or claims of clinical predictive accuracy. The activity-rhythm and at-home associations were exploratory and did not survive FDR correction. The observational, single-site design cannot establish temporal or causal mechanisms and examined only one medication and digital formulation. Several passive measures were proxies, and sensor and survey engagement declined over time, so later observations may reflect selective retention of more engaged participants. The mobility analyses added seven tests in 15 participants and 88 no-dose days and should be interpreted as checks on the accelerometer finding rather than independent results. Smartphone accelerometry measures device inactivity rather than sleep; rule-based and state-model rest durations correlated only moderately (r=0.36), and Fig 4 should be read as a statement about that construct, not about sleep and adherence.

**Fig 4.**
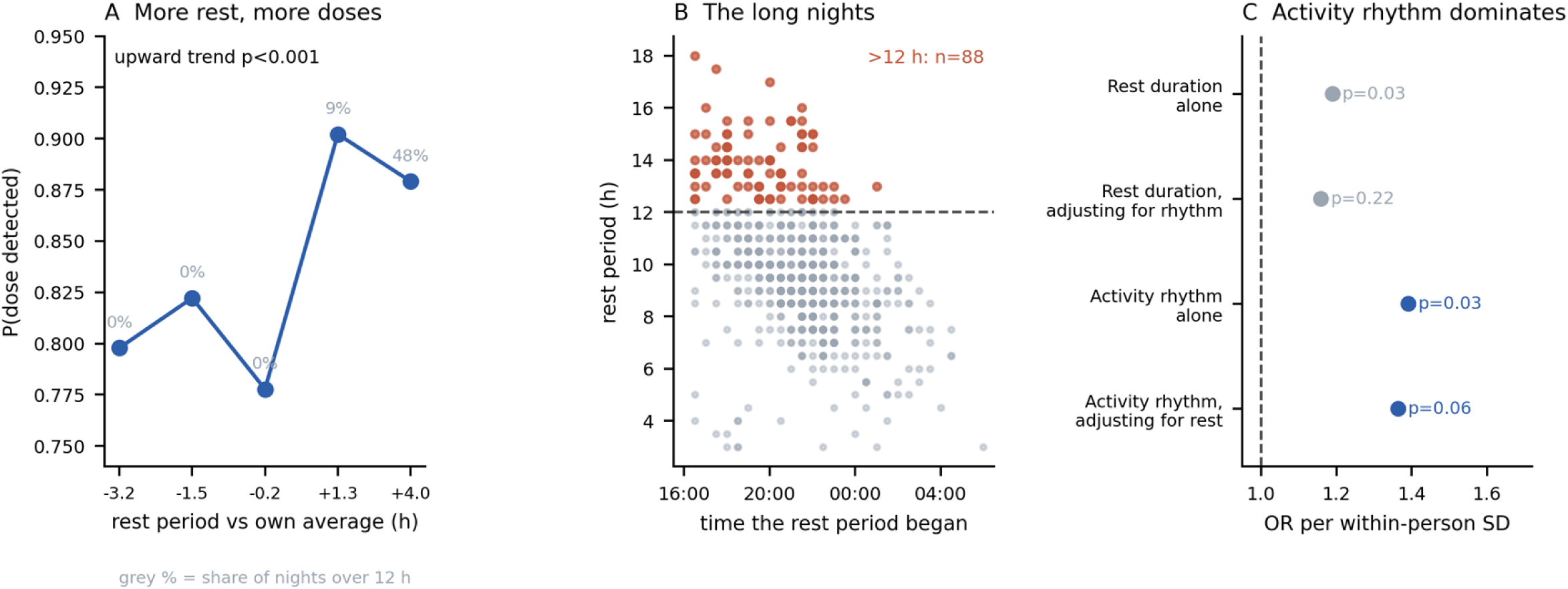
Longer-than-usual nighttime device inactivity and whether a dose was taken. These rest periods are derived from phone movement and screen use and record how long the phone was continuously still and unused; they measure device inactivity, not sleep. (A) Days were grouped into five equal bins according to how much longer or shorter the preceding rest period was than that participant’s own average. The probability of a detected dose rises across the bins. The rise begins in bins containing almost no nights longer than 12 hours (grey percentages), and adding a separate term for nights over 12 hours does not improve the model (p=0.29), so this is a gradual trend across the whole range rather than an effect of a few extreme nights. (B) Every recommended night plotted by when its rest period began and how long it lasted, with nights longer than 12 hours marked in orange. These are the nights that a filter based on plausible sleep duration would discard. They are shown separately to establish what they are: they begin in the evening and continue past midnight, 95% have complete 24-hour data, and the model still detects waking within them (mean efficiency 0.87). They are therefore valid recordings of a phone left untouched for a long period, not recording failures — and excluding them would remove genuine measurements of the construct rather than remove error. (C) Rest duration and activity-rhythm regularity entered in the same model. “Rest duration” is how long the phone was continuously inactive overnight; “activity rhythm” is how closely the day’s hour-by-hour movement resembled that participant’s own typical day. Activity rhythm remains associated with a detected dose after adjustment for rest duration (odds ratio 1.36, p=0.06), whereas rest duration does not survive adjustment for activity rhythm (1.16, p=0.22). The association is therefore carried by how typical the day was, with overnight rest duration acting as a further expression of the same consolidated, routine pattern.

In conclusion, interpretation of digital-medicine adherence requires explicit distinction between medication-taking and the ability to observe it. More than half of no-detection days in this study occurred without adequate patch wear, and conditioning on wear reduced apparent nonadherence from 32.3% to 18.1%. A three-state framework separates detected ingestion, adequately observed no-ingestion, and unobserved periods and provides a more transparent representation of adherence data from receiver-dependent systems. Secondary analyses suggested that medication-taking may also vary with participant-specific activity patterns and the immediate dosing environment, although these associations were exploratory. Digital adherence studies should therefore establish observation status before estimating adherence or interpreting behavioral correlates.

## MATERIALS AND METHODS

### Study design and participants

This single-site observational study analyzed adult outpatients with major depressive disorder (MDD) prescribed aripiprazole through the Abilify MyCite digital-medicine system. Twenty participants contributed objective ingestion and patch-contact data across 997 person-days collected between February 1st and October 1st 2024 (median 36.5 days per participant, range 16–106). Seventeen participants also contributed smartphone digital phenotyping and formed the primary behavioral analysis sample, providing 744 patch-worn person-days. Of these, 17 contributed accelerometry, 16 contributed EMA, 15 contributed GPS sufficient for location features, and 15 contributed all three. Participant demographic and baseline clinical characteristics are summarized in Table 1. The Beth Israel Deaconess Medical Center Institutional Review Board approved protocol 2023P-000628, and all participants provided written informed consent.

**Table 1.** Participant demographic characteristics.

| Characteristic | Participants (n = 20) |
| --- | --- |
| **Age, years** |  |
| Mean (SD) | 46.3 (18.36) |
| Missing | 15% (n = 3) |
| **Sex** |  |
| Male | 15% (n = 3) |
| Female | 70% (n = 14) |
| Other | 0% (n = 0) |
| Missing | 15% (n = 3) |
| **Race and ethnicity** |  |
| White | 75% (n = 15) |
| Black/African-American | 10% (n = 2) |
| Asian | 0% (n = 0) |
| Multiracial or Other | 0% (n = 0) |
| Hispanic | 0% (n = 0) |
| Missing | 15% (n = 3) |
| **Education** |  |
| Some high school or less | N/A |
| High school/some college or less | N/A |
| University or postgraduate degree | N/A |
| Missing | N/A |

### Abilify MyCite digital medicine system

Abilify MyCite combines aripiprazole with an ingestible event-marker sensor that transmits a signal on contact with gastric fluid to a wearable adhesive patch on the torso. The patch records ingestion time and relays the event to a smartphone application; with participant consent, ingestion records are available to the clinical team [8,9]. The patch also records an impedance-based skin-contact stream, which we used to distinguish no-detection days with adequate patch wear from days with insufficient wear.

### Smartphone digital phenotyping

Passive and active smartphone data were collected with mindLAMP, an open-source digital-phenotyping platform for Apple and Android devices [18]. Passive streams included accelerometry, screen-interaction state, and GPS-derived location, from which we derived physical activity, mobility, time at home, and location entropy. Screen-state and accelerometer data were also used to approximate sleep-related rest. Active data consisted of a brief daily EMA survey covering mood, anxiety, functioning, and sleep. All EMA and self-report measures analyzed here were collected through mindLAMP; MyCite in-app missed-dose survey responses were not included in the analytic dataset.

### Day-level adherence outcome

Because ingestion can be detected only while the patch is in contact with the skin, a day without a detected ingestion cannot be classified from ingestion data alone. We defined three daily states: (1) adherent, with at least one detected ingestion; (2) wear-supported no ingestion, with no detected ingestion but at least 6 hours of good skin contact; and (3) patch-off or indeterminate, with less than 6 hours of contact. The 6-hour threshold provided a substantial observation window while retaining sufficient days for analysis; sensitivity analyses at 1, 4, 8, and 12 hours yielded similar results. Total daily wear does not establish that the patch was worn at the exact dosing time, so the second state represents an adequately observed no-ingestion day rather than a confirmed missed dose. MyCite in-app missed-dose self-report was not used to adjudicate these states. Internal validation identified 12 days previously labeled nonadherent despite a recorded ingestion event, so day-level labels were rebuilt directly from the raw ingestion and skin-contact records.

### Behavioral features

Daily behavioral features were derived from accelerometry, screen interaction, GPS, and EMA. The prespecified primary family covered activity, mobility, screen use, and sleep-related rest. Activity-rhythm regularity, our main routine measure, was the correlation between each day’s hourly activity profile and that participant’s median profile across other study days; higher values indicate a more typical, not more active, day. A leave-one-day-out version produced the same estimate. We also examined a nine-feature routine/circadian family and a composite disruption index as exploratory analyses. Because dose timing was stable within participants (within-person SD ≈2 h), peri-dose analyses quantified time at home, movement, and screen use during the habitual dose hour ±2 h and during the preceding four hours. Features were centered on each participant’s mean and scaled by the pooled within-person SD, so estimates represent day-to-day deviations within a person. One within-person SD corresponds to 0.15 correlation units of activity-rhythm regularity and about 20 minutes spent at home during the habitual dose hour. GPS features followed prior work: location variance, normalized entropy, homestay, transition time, total distance, number of clusters, and radius of gyration. Two purpose-built measures tested the routine hypothesis: a location analogue of activity-rhythm regularity and time at home during the single habitual dose hour, estimated leave-one-day-out. Home was defined as the participant’s modal nighttime location, with at-home status within 300 m.

### Statistical analysis

Each feature was tested in a separate Bayesian mixed-effects logistic regression with participant random intercepts and within-and between-person decomposition; only within-person estimates are interpreted. Models adjusted for weekend, study week, sensor coverage, and EMA response and are reported as posterior odds ratios with 95% credible intervals. Variational approximation may underestimate posterior uncertainty, so multiplicity-corrected inference was based on frequentist sensitivity analyses rather than variational posterior intervals. Each association was re-estimated with generalized estimating equations (GEE), which yield population-averaged estimates; mixed-model and GEE results were compared for direction and interval overlap rather than numerical equality. GEE p values were corrected within each feature family using Benjamini-Hochberg (q<0.10). Correcting the variational p values instead would place the activity-rhythm association at q=0.06. The same features were tested against patch-off status and EMA nonresponse to assess general disengagement. For prospective prediction, penalized logistic regression used prior-day features and recent summaries with leave-one-subject-out cross-validation. Performance was evaluated using AUROC, precision-recall area, calibration, and a within-person permutation test and compared with covariate-only and first-week adherence base-rate models.

## Data Availability

The participant-level data underlying this study cannot be made publicly available because they contain sensitive health information and high-resolution digital phenotyping data, including GPS-derived location information, that may pose a risk of participant re-identification. Public sharing is therefore restricted by the informed consent under which the data were collected and by institutional ethics requirements. De-identified data necessary to reproduce the reported findings may be made available to qualified researchers upon reasonable request, subject to approval by the Beth Israel Deaconess Medical Center Institutional Review Board and any applicable data-use agreements. Requests for data access may be directed to the corresponding author.

## Acknowledgments

We thank the study participants and the Otsuka team for feedback on the manuscript.

